# Estimating the Cost of Typhoid Conjugate Vaccine Delivery in Ghana and the Democratic Republic of the Congo

**DOI:** 10.64898/2026.08.12.26360337

**Authors:** Kofi Akohene Mensah, Robert Lumbala, Yongha Hwang, Winthrop Morgan, Marie-France Phoba, Francis Opoku Agyapong, Michael Owusu, Jules Mbuyamba, Michael Owusu-Ansah, Thaint Thaint Thwe, Mohamadou Siribie, Jean-Paul Kumbukama, Pierre Cédric Khuwa, Hyonjin Jeon, Birkneh Tilahun Tadesse, Sampson Twumasi-Ankrah, Florian Marks, Octavie Lunguya, Ellis Owusu-Dabo, Jung-Seok Lee

## Abstract

Typhoid fever remains a significant burden in low- and middle-income countries (LMICs). The World Health Organization recommends incorporating typhoid conjugate vaccines (TCVs) into the routine immunization programs of typhoid-endemic countries. Although TCV has been shown to be safe, well tolerated, and effective, evidence on its delivery costs in African settings remains limited. This study provides economic evidence on the cost of implementing TCV catch-up campaigns.

This retrospective provider-perspective costing study evaluated TCV catch-up vaccination campaigns conducted in the Asante-Akim North District of Ghana and the Kisantu Health Zone of the Democratic Republic of the Congo (DRC). The campaigns targeted children aged 9 months to 15 years. An incremental costing approach was used, and a Microsoft Excel-based tool was developed to estimate costs.

The total number of vaccinated individuals was 54,814; 10,052 in Ghana and 44,762 in the DRC. The financial cost per fully immunized person (FIP), including vaccine and vaccination supply costs, was estimated at US$ 5.78 in Ghana and US$ 5.47 in the DRC. The corresponding economic costs were estimated at US$ 6.09 in Ghana and US$ 5.89 in the DRC. Vaccine procurement and vaccination supplies represented the largest cost component, accounting for US$ 2.76 per FIP in Ghana and US$ 2.39 per FIP in the DRC, followed by service delivery and service delivery support activities.

This study provides empirical estimates of the financial and economic costs of TCV catch-up campaigns in Ghana and the DRC. These findings provide country-specific evidence to inform planning, budgeting, economic evaluation, and policy decisions regarding future TCV introduction in typhoid-endemic settings.

## INTRODUCTION

Typhoid fever remains a significant public health burden in low- and middle-income countries (LMICs), with an estimated 11–21 million cases occurring annually worldwide [1]. Although typhoid fever can affect all age groups, its incidence is higher among preschool- and school-aged children compared to older individuals [2]. Studies on typhoid fever incidence identify south-central Asia, southeast Asia, and southern Africa as high-incidence regions, particularly in areas with limited access to improved water quality and inadequate sanitation [3].

Ghana and the Democratic Republic of the Congo (DRC) are typhoid-endemic countries. In Ghana, the 2023 Global Burden of Disease (GBD) study estimated an incidence of 62 typhoid cases per 100,000 population, with more than 70% among children aged 14 years or younger. An estimated 332 typhoid deaths, of which 77% were in the same group [4]. In the DRC, multiple large-scale typhoid outbreaks over the past decade were observed, highlighting the continued public health importance of typhoid fever in the country [5]. Hospital surveillance data from the DRC indicate that typhoid is frequently identified in children, of whom 72% were below the age of ten [6]. Consistent with these observations, the GBD 2023 study estimated approximately 252 typhoid cases per 100,000 population, with more than 60% of cases occurring among children aged 14 years or younger. There were 3,295 typhoid deaths [4].

The World Health Organization (WHO) Strategic Advisory Group of Experts (SAGE) on Immunization recommended in October 2017 the use of typhoid conjugate vaccines (TCVs) in the routine immunization programs of typhoid-endemic countries [7] and GAVI added TCV into its battery of subsidized vaccines and put US$ 85 million aside to support the introduction of TCVs in GAVI-eligible countries [8]. To date, nine GAVI-eligible countries have introduced TCV into their routine immunization schedules [9], while additional countries have implemented TCV through targeted vaccination programs. During the study period, both Ghana and the DRC were GAVI-eligible countries [10]. As more countries consider TCV introduction, there is an increasing need for evidence on the economic burden of typhoid fever and the cost-effectiveness of alternative TCV introduction strategies, including introducing TCVs into routine immunization, along with catch-up campaigns for children up to 15 years of age.

A single dose of TCV provides protection for at least four years among children aged 9 months to 12 years and has demonstrated effectiveness across age groups, including children younger than two years [11]. While this evidence supports the potential public health value of TCV introduction, implementation decisions also require reliable estimates of the resources and costs associated with vaccine delivery. Such evidence remains limited for TCV campaigns in African settings. Real-world delivery cost data are essential for estimating the budgetary and economic implications of TCV introduction, informing cost-effectiveness analyses, and supporting evidence-based decision-making regarding TCV adoption in high-burden settings.

From the provider perspective, this study retrospectively estimated the incremental financial and economic costs of TCV catch-up vaccination campaigns implemented in Ghana and the DRC to inform future TCV planning, budgeting, and implementation.

## METHODS

### Study sites and Vaccination campaigns

This study retrospectively estimated the costs of two previously implemented TCV catch-up vaccination campaigns in Ghana and the DRC. The campaigns were conducted in the Asante-Akim North District of Ghana between August 2021 and March 2022, and Kisantu Health Zone of the DRC between February 2022 and May 2023. The target population for TCV was children and adolescents aged 9 months to 15 years. One dose was required for a Fully Immunized Person (FIP).

In the Asante-Akim North District, the study focused on five areas: Agogo, Ananekrom, Nyinamponase, Hwidiem, and Juansa. The eligible target population was estimated at 22,539 individuals. Half of the participants received TYPBAR-TCV®, a Vi-polysaccharide conjugated to a tetanus-toxoid protein carrier (Vi-TT), while the other half received a control vaccine, MenAfriVac (group A meningococcal polysaccharide-tetanus toxoid conjugate vaccine [MCV-A]).

In the Kisantu Health Zone of the DRC, 17 areas were included in the study, including Nkandu, Nzeza Nlandu and Gare, among others. The eligible target population was estimated at 85,860 individuals. A mass vaccination campaign using TYPBAR-TCV^®^ was conducted, and no control-vaccine group was included. The population for each area in this study is detailed in Annex Tables S1 – S3.

### Study design

The costing study took the provider’s perspective, estimating costs for planning and conducting a TCV catch-up campaign targeting children. The study used an ‘ingredients-based’ costing methodology, i.e., all personnel, allowances, consumables, and other direct costs were identified. Resource quantities were multiplied by their corresponding unit costs to estimate activity-specific costs. Activity-specific costs were multiplied by the frequency of implementation to estimate total costs.

An incremental costing approach was used. Only those activities and resources used specifically for TCV vaccination-related activities were costed. Shared resources, such as personnel time, transportation, and cold chain equipment, were allocated according to the proportion attributable to TCV-related activities, as determined from project financial records and interviews with project managers. In cases where detailed activity cost data was unavailable, aggregated expenditure records were used.

The study calculated financial costs (i.e., monetary outlays) and economic costs (i.e., financial costs plus the value of existing, in-kind, and donated resources). The estimated results are presented in US dollars; costs were adjusted to 2023 price levels using country-specific GDP deflators and subsequently converted to US dollars using the corresponding exchange rates [12, 13]. Cost items and activities were categorized as follows in Tables 1 and 2.

**Table 1.** Cost Categories for TCV Costing.

| **Cost Category** | **Includes** |
| --- | --- |
| Vaccines and Vaccination Supplies | Vaccine, auto-disable syringes with needles, safety boxes. |
| Personnel | Project personnel, government personnel |
| Allowances | Transport allowance, holiday allowance, overtime allowance, meal allowance, etc. |
| Consumables | Gloves, cleaning supplies, stationery, flyers, etc. |
| Other Direct Costs | Refreshments, meals, hall rental, public announcement packages, professional services such as facilitators, fuel, etc. |
| Cold Chain Equipment | Any cold chain equipment with one or more useful life years purchased for use by the TCV vaccination activities (allocated according to use proportion) |

**Table 2.**
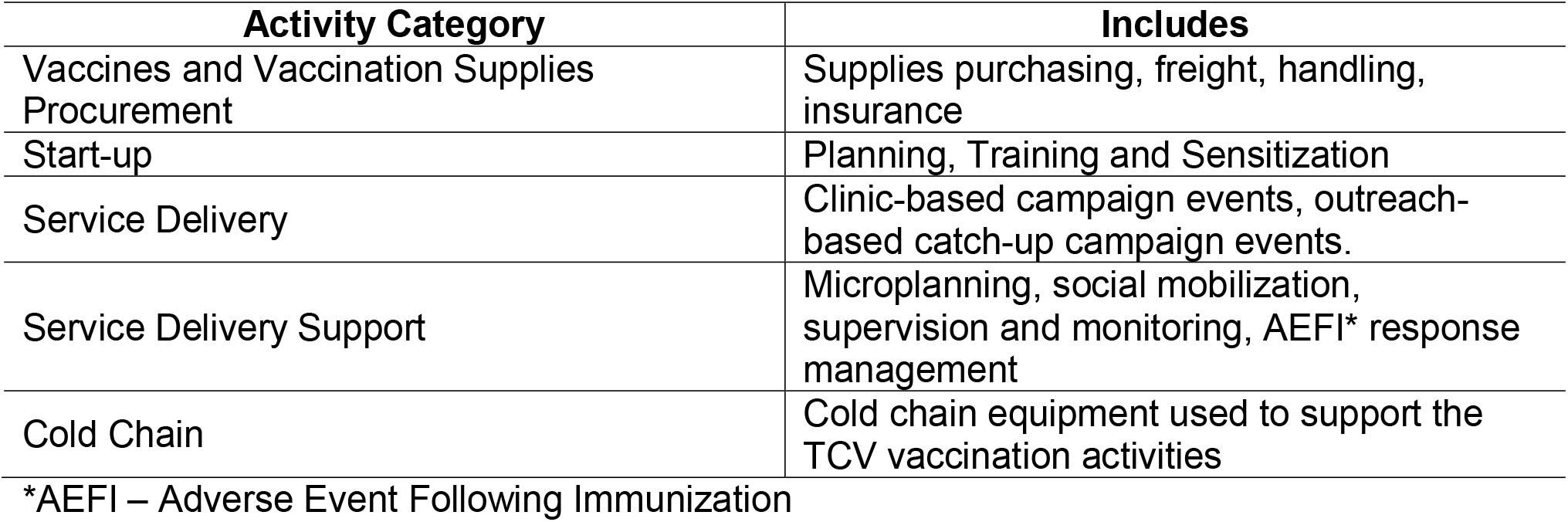
Activity Categories for TCV Costing.

| **Activity Category** | **Includes** |
| --- | --- |
| Vaccines and Vaccination Supplies Procurement | Supplies purchasing, freight, handling, insurance |
| Start-up | Planning, Training and Sensitization |
| Service Delivery | Clinic-based campaign events, outreach-based catch-up campaign events. |
| Service Delivery Support | Microplanning, social mobilization, supervision and monitoring, AEFI* response management |
| Cold Chain | Cold chain equipment used to support the TCV vaccination activities |

\*AEFI – Adverse Event Following Immunization

The vaccination campaign activities were categorized into five major cost categories (Table 2). Within these categories, program activities were further grouped into nine activity groups, with the detailed activities included in each group presented in Annex Table S4. Transportation-related expenses were allocated to the relevant activity categories, including service delivery and service delivery support, as appropriate.

Start-up costs associated with vaccine introduction, including sensitization, training, and introduction planning, were assumed to have a useful life of five years. It was further assumed that the cold chain equipment purchased to support the campaign had useful lives between three and ten years, according to the manufacturer. However, a detailed breakdown of the cold chain equipment costs in the DRC was not available, so the useful life was assumed to be the average of Ghana’s. Start-up and cold chain equipment costs were converted into annual equivalent costs over their assumed useful lives using a 3% discount rate. Annual equivalent costs were allocated in one-year increments according to the duration of campaign implementation. This approach was considered appropriate because the benefits of start-up activities and the use of cold chain equipment extend beyond the period of vaccine administration, and their utilization cannot be precisely attributed to individual months. In this study, the cost of wasted vaccines and injection syringes was excluded. Output included the total cost of the campaign and the cost per FIP.

### Data collection

A Microsoft Excel-based tool was developed to estimate the vaccination campaign costs using a micro-costing approach, in which each resource used was identified, quantified, and assigned a unit cost before calculating total costs. Building costs and project overhead costs were excluded from the analysis. Data was collected from the project office financial records and through interviews with project managers. Data collection was conducted by trained graduate research assistants from the Kumasi University for Science and Technology (KNUST) in Ghana and by the Finance Manager of the Institut National de Recherche Biomédicale (INRB) in the DRC. They received training in cost analysis and cost data collection and were supervised and monitored throughout the data collection period. Interview-derived information was cross-checked against available financial and programmatic records, and any inconsistencies were clarified with the relevant project personnel.

Data was entered into a Microsoft Excel-based workbook, which organized detailed costs into activities and categories and then calculated outputs and costs.

### Ethical statement

The delivery cost estimation of TCV in both countries presented here is part of research protocols *Typhoid Conjugate Vaccine Effectiveness in Ghana* (TyVEGHA) and *An open-label effectiveness study of a typhoid conjugate vaccine in Kisantu, Democratic Republic of the Congo* (TyVECO). These studies were approved by the Institutional Review Board (IRB) of the International Vaccine Institute (IVI), as well as by the Ghana Health Service Ethics Review Committee, the Committee on Human Research, Publications and Ethics of Kwame Nkrumah University of Science and Technology (CHRPE-KNUST), and the Ghana Food and Drugs Authority (FDA) in Ghana, and the Ethics Committee of the School of Public Health at the University of Kinshasa in the DRC.

## RESULTS

### Campaign implementation and coverage rate

The campaign vaccinated 54,814 persons: 10,052 in Ghana and 44,762 in the DRC between the ages of 9 months and 15 years. In Ghana, a total of 192 vaccination events occurred during the campaign. Of these, 178 were clinic-based events, during which 8,904 persons were vaccinated, and 14 were outreach events, during which 1,148 persons were vaccinated. Following the clinic-based service delivery phase of the campaign, the outreach campaign was conducted in the peripheral areas of the largest area (Agogo, target population 6,184). An additional seven were held in the smallest area (Hwidiem, target population 724). The total coverage rate was estimated to be 89.2% (Figure 1). Vaccination coverage exceeded 100% in Hwidiem, Ananekrom, and Nyinamponase because the coverage denominator was based on the originally allocated TCV target population. Under the Ghanaian vaccination strategy, approximately half of the eligible population was allocated to receive TCV and the other half to receive MCV-A. During implementation, the number of individuals who received TCV exceeded the originally allocated TCV target in these areas.

**Figure 1.**
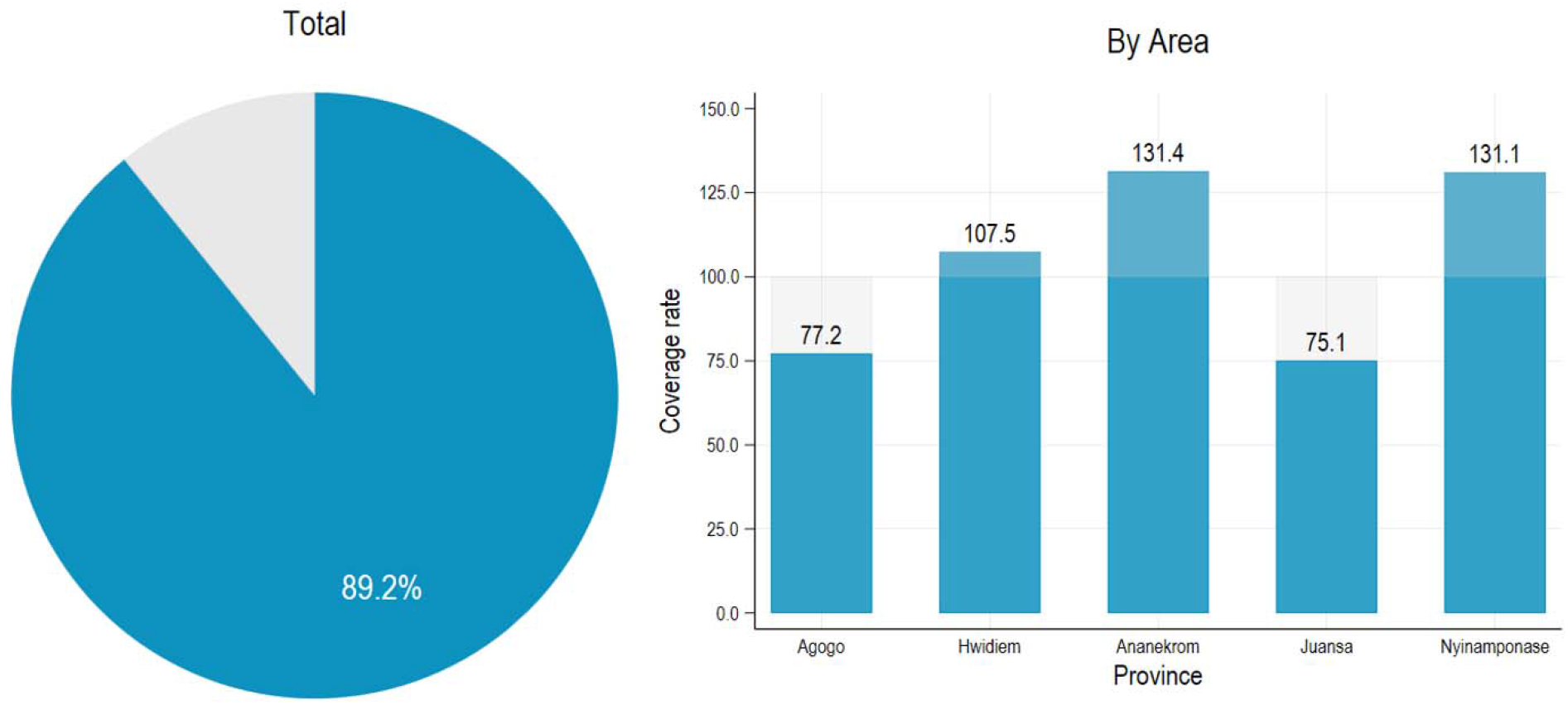
Vaccination Coverage Achieved - Ghana

In the DRC, vaccination events occurred in clinics, and there were no outreach events. The vaccination events were conducted in 36 health facilities in 17 areas for an average of 27.9 days and up to 93 days, during which 44,762 persons were vaccinated. The total coverage rate was estimated to be 52.1% (Figure 2). Area-specific vaccination coverage is presented in Annex Tables S2 and S3.

**Figure 2.**
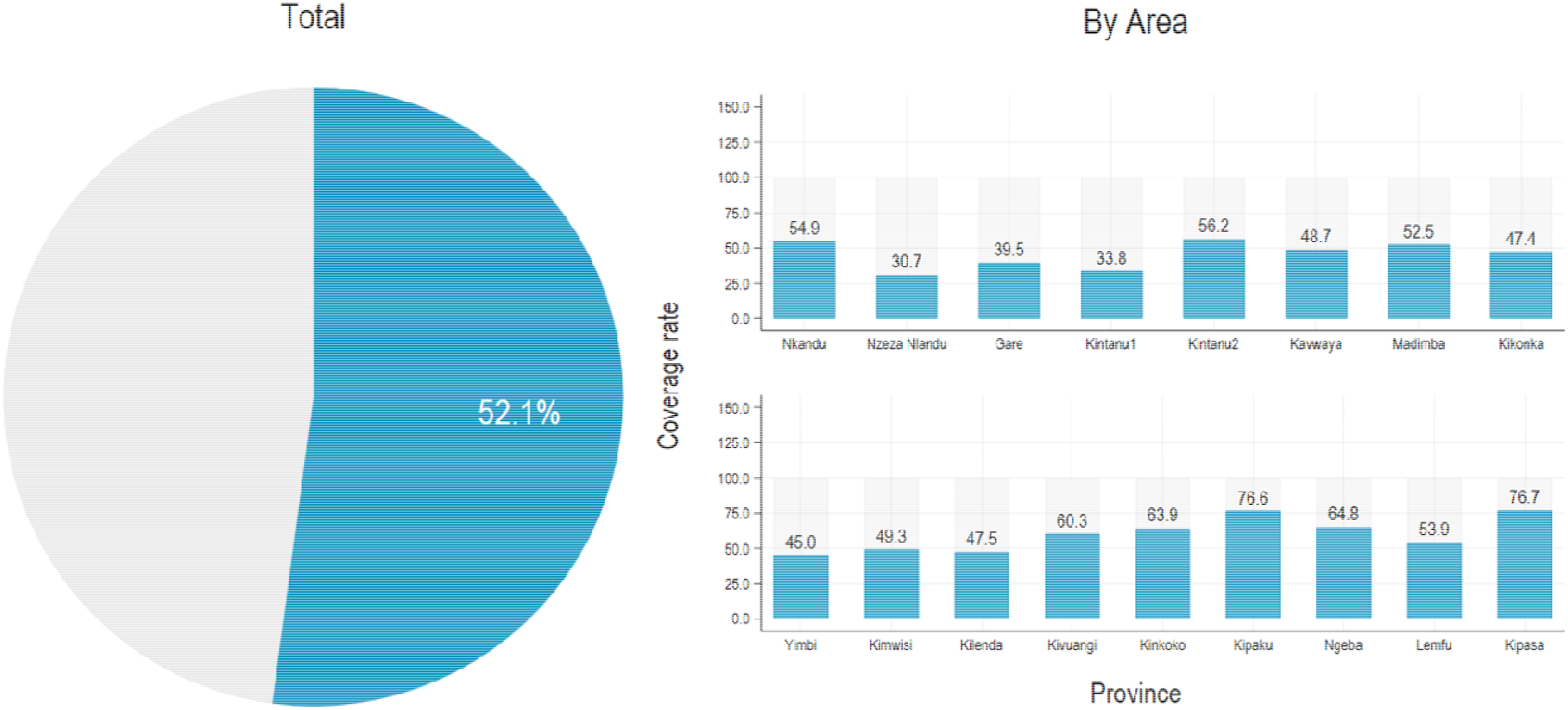
Vaccination Coverage Achieved - DRC

### Cost of the vaccination campaign

The total vaccination campaign cost was estimated at US$ 58,144 in financial costs and US$ 61,259 in economic costs in Ghana (Figure 3). In the DRC, the total cost was estimated to be US$ 244,708 in financial costs and US$ 263,551 in economic costs.

**Figure 3.**
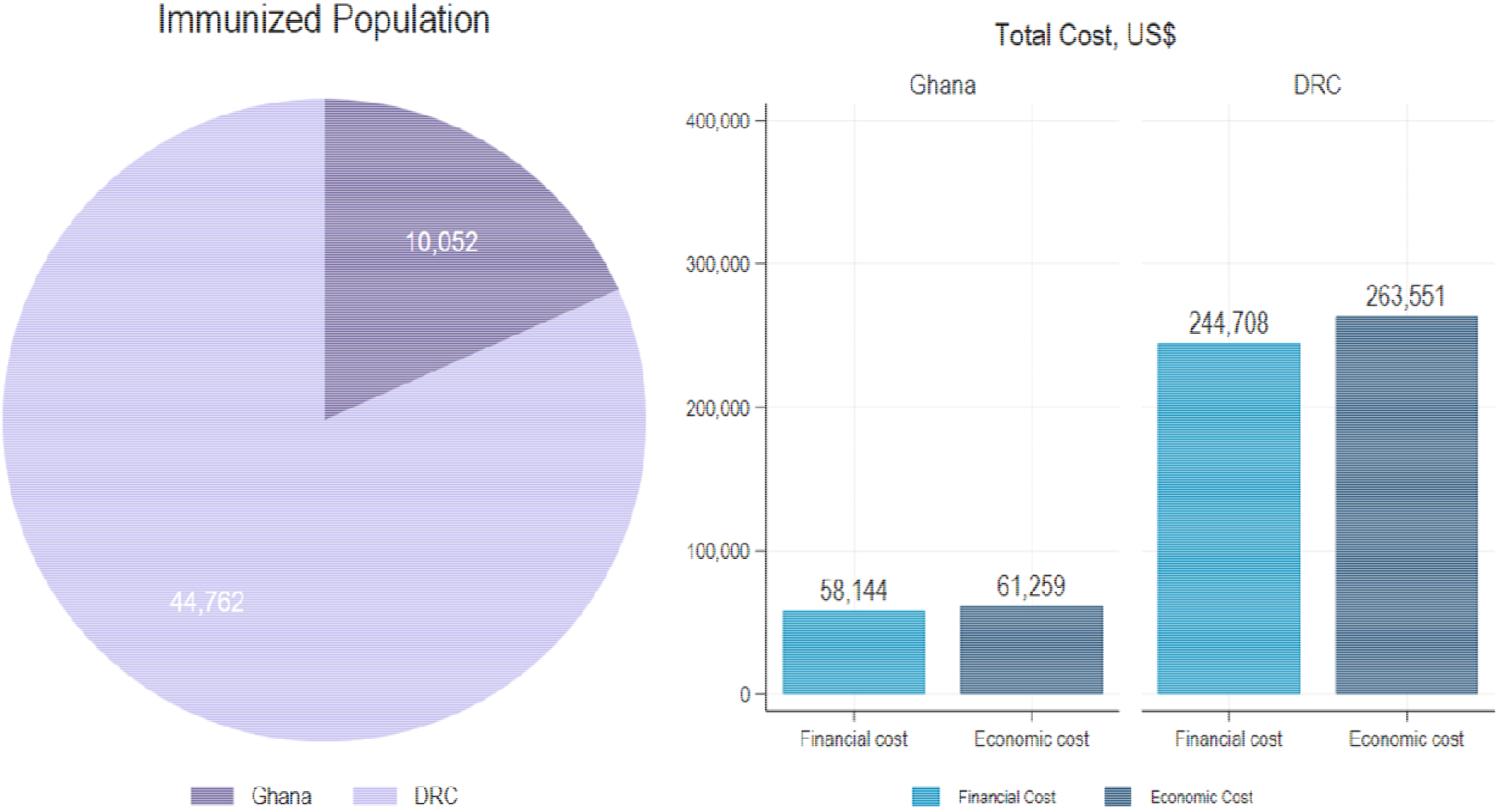
Total Vaccinated Population and Campaign Costs

The financial and economic costs per FIP for the TCV campaign by activity are shown in Table 3. The cost per FIP was estimated at US$ 5.78 in financial costs and US$ 6.09 in economic costs in Ghana. In the DRC, the cost per FIP was estimated at US$ 5.47 in financial costs and US$ 5.89 in economic costs. Across all categories, the highest financial and economic cost per FIP was incurred for purchase of vaccines and vaccination supplies. Excluding vaccine procurement costs, service delivery and service delivery support accounted for the largest share of campaign delivery costs in both countries.

**Table 3.**
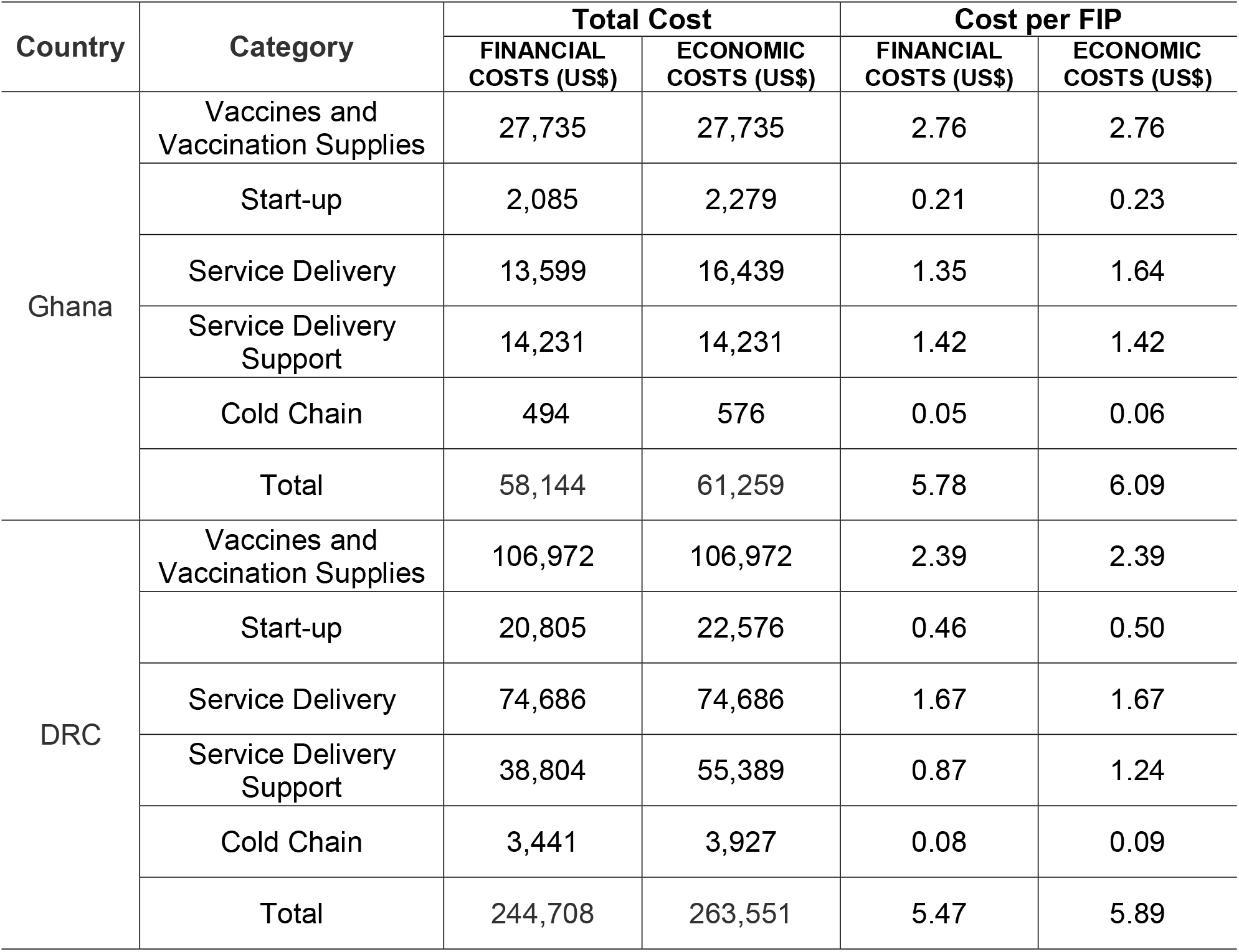
Costs of TCV Campaign.

| Country | Category | Total Cost |  | Cost per FIP |  |
| --- | --- | --- | --- | --- | --- |
| | | FINANCIAL COSTS (US\$) | ECONOMIC COSTS (US\$) | FINANCIAL COSTS (US\$) | ECONOMIC COSTS (US\$) |
| Ghana | Vaccines and Vaccination Supplies | 27,735 | 27,735 | 2.76 | 2.76 |
|  | Start-up | 2,085 | 2,279 | 0.21 | 0.23 |
|  | Service Delivery | 13,599 | 16,439 | 1.35 | 1.64 |
|  | Service Delivery Support | 14,231 | 14,231 | 1.42 | 1.42 |
|  | Cold Chain | 494 | 576 | 0.05 | 0.06 |
|  | Total | 58,144 | 61,259 | 5.78 | 6.09 |
| DRC | Vaccines and Vaccination Supplies | 106,972 | 106,972 | 2.39 | 2.39 |
|  | Start-up | 20,805 | 22,576 | 0.46 | 0.50 |
|  | Service Delivery | 74,686 | 74,686 | 1.67 | 1.67 |
|  | Service Delivery Support | 38,804 | 55,389 | 0.87 | 1.24 |
|  | Cold Chain | 3,441 | 3,927 | 0.08 | 0.09 |
|  | Total | 244,708 | 263,551 | 5.47 | 5.89 |

### Vaccines and vaccination supplies cost

Vaccines were purchased from an international source at a market price of US$ 2.00 per dose. After adding freight, handling, and insurance costs, the vaccine cost was estimated at US$2.45 per FIP in Ghana and US$2.25 per FIP in the DRC (Table 4). Auto-disable syringes and needles were purchased at a market price of US$ 0.13 each in Ghana and US$ 0.12 each in the DRC. Safety boxes were purchased at an average price of US$ 18.12 in Ghana and US$ 2.20 in the DRC. Overall, vaccine and vaccination supply costs, including injection syringes and safety boxes, were estimated at US$ 2.76 per FIP in Ghana and US$ 2.39 per FIP in the DRC.

**Table 4.** Costs of Vaccines and Injectable Supplies.

| Cost Category | Ghana |  | DRC |  |
| --- | --- | --- | --- | --- |
| | Total Cost (US\$) | Cost per FIP (US\$) | Total Cost (US\$) | Cost per FIP (US\$) |
| Vaccine | 24,585 | 2.45 | 100,616 | 2.25 |
| Injection Syringe | 1,332 | 0.13 | 5,371 | 0.12 |
| Safety Box | 1,828 | 0.18 | 985 | 0.02 |
| Total | 27,735 | 2.76 | 106,972 | 2.39 |

### Start-up (Introduction) costs

Since TCV was a new vaccine in the area, start-up (Introduction) activities were included in the campaign in the start-up phase, and consisted of planning and preparation, training, and sensitization activities.

Start-up costs were estimated at US$ 0.23 per FIP in Ghana and US$ 0.50 per FIP in the DRC (Table 5). Training represented the largest component of start-up costs in both countries.

**Table 5.** Startup Costs.

| Country | Category | Total Cost |  | Cost per FIP |  |
| --- | --- | --- | --- | --- | --- |
| | | FINANCIAL COSTS (US\$) | ECONOMIC COSTS (US\$) | FINANCIAL COSTS (US\$) | ECONOMIC COSTS (US\$) |
| Ghana | Planning and Preparation | 467 | 515 | 0.05 | 0.05 |
|  | Training | 1,158 | 1,264 | 0.12 | 0.13 |
|  | Sensitization | 458 | 500 | 0.05 | 0.05 |
|  | Total | 2,085 | 2,279 | 0.21 | 0.23 |
| DRC | Planning and Preparation | 5,106 | 5,493 | 0.11 | 0.12 |
|  | Training | 12,436 | 13,573 | 0.28 | 0.30 |
|  | Sensitization | 3,263 | 3,511 | 0.07 | 0.08 |
|  | Total | 20,805 | 22,576 | 0.46 | 0.50 |

### Service delivery costs

Service Delivery includes all vaccination-related activities. In Ghana, the economic cost per FIP was estimated at US$ 1.77 for clinic-based events and US$ 0.56 for outreach events (Table 6). In the DRC, all vaccination activities were conducted through clinic-based campaigns, and the economic cost per FIP was estimated at US$ 1.67. No outreach-based vaccination activities were implemented in the DRC.

**Table 6.** Service Delivery Costs.

| Country | Category | Total Cost |  | Cost per FIP |  |
| --- | --- | --- | --- | --- | --- |
| | | FINANCIAL COSTS (US\$) | ECONOMIC COSTS (US\$) | FINANCIAL COSTS (US\$) | ECONOMIC COSTS (US\$) |
| Ghana | Clinic-based Campaign | 13,207 | 15,793 | 1.48 | 1.77 |
|  | Outreach-based Catch-up Campaign | 392 | 646 | 0.34 | 0.56 |
|  | Total | 13,599 | 16,439 | 1.35 | 1.64 |
| DRC | Clinic-based Campaign | 74,686 | 74,686 | 1.67 | 1.67 |
|  | Outreach-based Catch-up Campaign | - | - | - | - |
|  | Total | 74,686 | 74,686 | 1.67 | 1.67 |

\* Cost per FIP was calculated using 8,904 vaccinated individuals for clinic-based activities and 1,148 vaccinated individuals for outreach activities in Ghana

### Service delivery support activity costs

Several activities were carried out before and during the campaign service delivery events to ensure that they ran smoothly. Service delivery support activities included microplanning, social mobilization, supervision and monitoring, and AEFI response. Service delivery support costs were US$ 1.42 per FIP in Ghana and US$ 1.24 per FIP in the DRC. Supervision and monitoring accounted for the largest share in Ghana, whereas microplanning represented the largest component in the DRC (Table 7). No AEFI-related costs were recorded in the DRC.

**Table 7.**
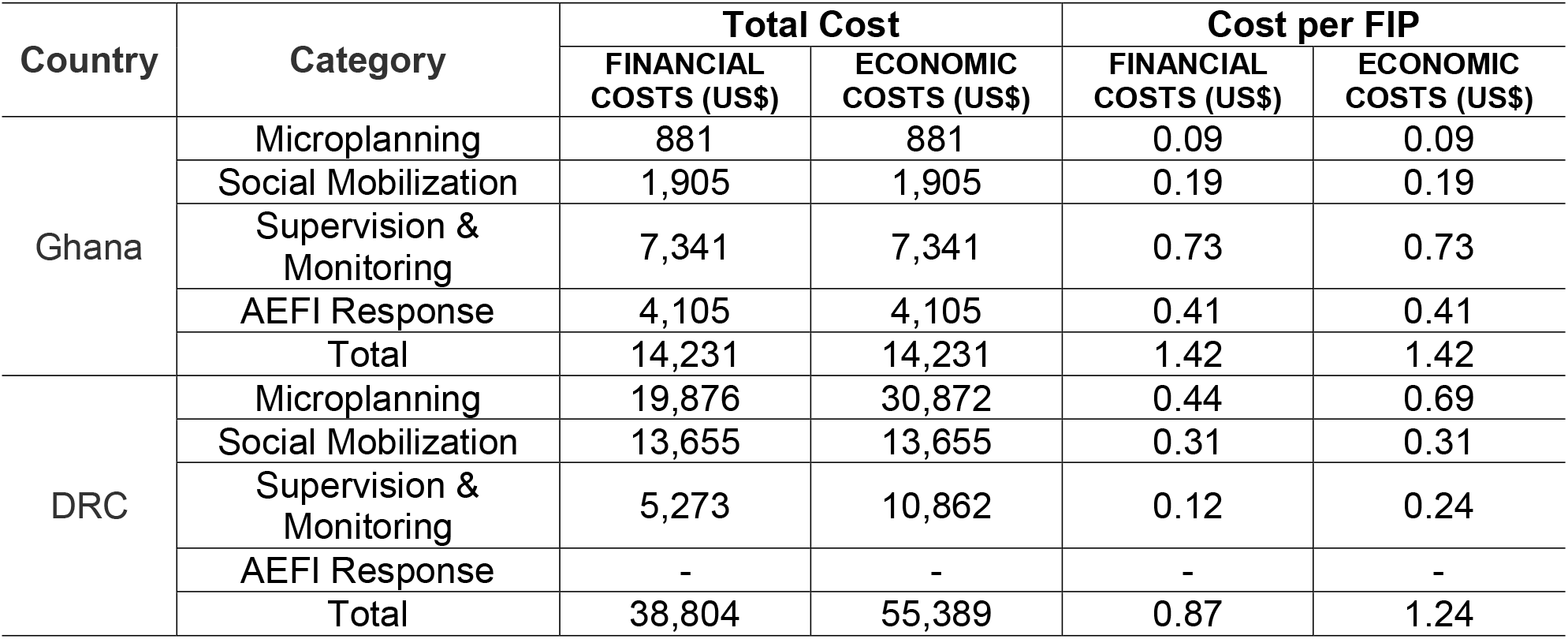
Service Delivery Support Costs.

### Cold chain costs

The total cost of cold chain equipment purchased in Ghana was US$ 4,699, and a detailed list of the equipment is provided in Annex Table S5. In the DRC, total costs related to cold chain equipment were reported at US$ 14,145. The annualized economic cost of cold chain equipment was estimated at US$ 0.057 per FIP in Ghana and US$ 0.088 per FIP in the DRC.

## DISCUSSION

This study estimated the financial and economic costs of delivering a single-dose TCV campaign in Ghana and the DRC. A total of 54,814 children and adolescents were vaccinated across both countries.

The average cost per FIP for the vaccination campaign was estimated at US$ 5.78 for financial costs and US$ 6.09 for economic costs in Ghana. For the DRC, the corresponding costs were estimated at US$ 5.47 and US$ 5.89, respectively. Excluding vaccine and vaccination supply costs, the economic cost was estimated to be US$ 3.34 in Ghana and US$ 3.50 in the DRC.

Since GAVI funding for TCV became available in 2018, several countries, including Nepal, Pakistan, Liberia, and Zimbabwe, have introduced TCV, with additional countries subsequently adopting the vaccine [14]. Excluding vaccine and vaccination supply costs, the cost per FIP estimated in Ghana and the DRC was higher than those reported in India (US$ 0.38 financial, US$ 1.49 economic), Zimbabwe (US$ 0.79 financial), Malawi (US$ 0.49 financial, US$ 0.84 economic), and Burkina Faso (US$ 0.47 financial; US$ 2.16 economic) [15-18]. The higher unit delivery costs observed in Ghana and the DRC may reflect variations in campaign design, costing methodology, and implementation context. One important explanation is the relatively small scale of the campaigns, which vaccinated approximately 113,000, 319,000, more than 7 million, and more than 10 million individuals in India, Zimbabwe, Malawi, and Burkina Faso, respectively. Larger campaigns are generally able to spread fixed costs over a greater number of vaccine recipients, resulting in lower unit delivery costs. Consistent with this finding, previous reviews of vaccine delivery costs in LMICs reported average delivery costs ranging from US$ 1.87 to US$ 2.47 per dose, although the reported ranges extended up to US$ 11.31 per dose [19-21]. Accordingly, although the delivery costs estimated in this study were higher than the reported averages, they fell within the range observed across LMICs.

These findings have important implications for future TCV introduction. Delivery costs varied substantially across settings, suggesting that countries should incorporate context-specific delivery costs into budgeting and economic evaluations rather than relying on a single unit cost estimate. Generating country-specific delivery cost evidence during vaccine introduction can support more realistic resource planning and improve the efficiency of future TCV implementation.

The study had several limitations. First, vaccination strategies differed between Ghana and the DRC. In Ghana, half of the eligible population was targeted for MCV-A vaccination and the other half for TCV vaccination, whereas in the DRC, the entire eligible population was targeted for TCV vaccination. Differences in delivery cost estimates may also have been associated with campaign scale, delivery modalities, resource prices, and the allocation of shared personnel and logistical resources. However, because only two campaigns were evaluated and comparable site-level explanatory variables were not systematically collected, the contribution of individual factors could not be quantified. The cost differences between the two countries should therefore be interpreted cautiously rather than attributed to any single aspect of campaign implementation.

Second, both campaigns were implemented within research projects rather than national immunization programs. Planning, supervision, monitoring, and staffing arrangements may differ from those used in routine or nationwide implementation. Conversely, larger national campaigns may achieve economies of scale, although they may also require additional coordination, logistics, and health-system resources. The estimates should therefore be transferred to other settings with consideration of local delivery structures and resource prices.

Third, during the data collection process, we did not conduct interviews with all staff involved in the vaccination campaigns. Cost information was obtained from key personnel responsible for campaign implementation and financial management. However, these individuals may not have directly observed all activities performed by others (e.g., time spent by each person in the activity).

Lastly, costs were estimated from research-supported implementation settings and may not fully reflect the costs of programmatically implemented TCV catch-up campaigns. Although efforts were made to exclude research-specific activities and allocate only vaccination-related costs, some operational characteristics of the study setting may differ from those of routine campaign implementation. In addition, building and overhead costs were not included in the cost analysis. As a result, the estimated costs may not fully capture all resources associated with vaccination campaign implementation and may not be directly comparable with studies that included these cost components. Therefore, these cost estimates should be interpreted with caution when extrapolating to large-scale TCV vaccination campaigns.

## CONCLUSION

In this study, the financial and economic costs of delivering TCV catch-up campaigns were estimated in Ghana and the DRC. These findings provide important evidence for estimating resource requirements and informing economic evaluations of TCV introduction. As additional countries consider introducing TCV as part of typhoid prevention and control strategies, the cost estimates generated in this study may support national and global decision-making regarding the planning and implementation of TCV vaccination programs.

## Supporting information

annex

## Data Availability

Aggregated data may be made available by the corresponding author upon reasonable request and subject to the conditions of the ethical approval.

