## Supplementary material for "Estimating the Cost of Typhoid Conjugate Vaccine Delivery in Ghana and the Democratic Republic of the Congo": annex

**Appendix 1.** Vaccination Coverage Rate Estimates

**Table S1. Eligible Target Population in Ghana**

| **Area** | **Eligible Population** |
| --- | --- |
| Agogo | 12,368 |
| Hwidiem | 1,447 |
| Ananekrom | 1,585 |
| Juansa | 4,357 |
| Nyinamponase | 2,782 |
| Total | 22,539 |

**Table S2. Vaccination Coverage Achieved in Ghana**

| **Area** | **Target Population for TCV** | **Immunized** | **Coverage** |
| --- | --- | --- | --- |
| Agogo | 6,184 | 4,773 | 77.2% |
| Hwidiem | 724 | 778 | 107.5% |
| Ananekrom | 793 | 1,042 | 131.4% |
| Juansa | 2,179 | 1,636 | 75.1% |
| Nyinamponase | 1,391 | 1,823 | 131.1% |
| Total | 11,270 | 10,052 | 89.2% |

**Table S3. Eligible Target Population and Vaccination Coverage Achieved in the DRC**

| **Area** | **Eligible Population** | **Immunized** | **Coverage** |
| --- | --- | --- | --- |
| Nkandu | 12,447 | 6,830 | 54.9% |
| Nzeza Nlandu | 7,276 | 2,232 | 30.7% |
| Gare | 2,976 | 1,177 | 39.5% |
| Kintanu 1 | 4,392 | 1,484 | 33.8% |
| Kintanu 2 | 8,499 | 4,777 | 56.2% |
| Kavwaya | 5,391 | 2,624 | 48.7% |
| Madimba | 4,876 | 2,561 | 52.5% |
| Kikonka | 7,829 | 3,709 | 47.4% |
| Yimbi | 1,929 | 869 | 45.0% |
| Kimuisi | 4,383 | 2,163 | 49.3% |
| Kilenda | 1,734 | 823 | 47.5% |
| Kivuangi | 2,405 | 1,450 | 60.3% |
| Kinkoko | 1,693 | 1,082 | 63.9% |
| Kipako | 4,547 | 3,483 | 76.6% |
| Ngeba | 8,084 | 5,240 | 64.8% |
| Lemfu | 6,217 | 3,351 | 53.9% |
| Kipasa | 1,182 | 907 | 76.7% |
| Total | 85,860 | 44,762 | 52.1% |

**Appendix 2.** Program activities included in cost analysis of the typhoid conjugate vaccine (TCV) campaign

**Table S4. Activity Groups for TCV Costing**

| **Activity Group** | **Includes** |
| --- | --- |
| Vaccines and Vaccination Supplies Procurement | - Vaccines and Vaccination Supplies purchasing - Shipping - Insurance |
| Planning and Preparation Activities | - Project-level planning and preparation for the campaign |
| Microplanning Activities | - Project Coordinator Pre-Campaign Supervisory Visit - HQ Microplanning |
| Training Activities | - Field staff campaign preparation training - Clinical Practice certification training |
| Sensitization Activities | - Briefings for traditional leaders - Briefings for local government authorities - Briefings for local government medical personnel |
| Social Mobilization Activities | - Community Announcements - One Day of Community Meetings (Church/Mosque/etc) in a Catchment Area - Flyer distribution - Information Centre Announcements |
| Service Delivery Activities | - Clinic-based Vaccination Campaign Event - Outreach Vaccination Campaign Event |
| Supervision and Monitoring Activities | - Pre-Campaign Preparation Supervision - Campaign Supervision (Vaccination Days) |
| AEFI Activities | - Hospital-based AEFI Response (paid by provider, including transport to and from hospital) |

**Appendix 3.** Cold chain equipment purchased in Ghana

**Table S5. Cold Chain Initial Investment in Ghana**

| **Equipment** | **USEFUL LIFE** | **FINANCIAL**  **COSTS (US$)** |
| --- | --- | --- |
| MainsRef Coolfinity IceVolt300P E003/122 | 10 | 392 |
| Fridge Thermometer | 10 | 46 |
| Roch RFR-170DT-J Double Door Refrigerator - 138 Litres Silver | 10 | 263 |
| Rainbow refrigerator 370ix double door | 10 | 402 |
| Vaccine Carrier Bags | 10 | 186 |
| Cold Box Small | 10 | 2,128 |
| Cold Box Large | 10 | 1,176 |
| Water pack B medical WP 0.3L (pack of 40) | 3 | 105 |
| Total Initial Investment |  | 4,699 |

* For the Ghana vaccination campaign, a refrigerator with a storage capacity of 241 liters was purchased and used to store both TCV and MenAfriVac vaccines. Based on the proportion of storage volume allocated to TCV (34 of 241 liters; 14%), 14% of the refrigerator cost was attributed to TCV. The project also purchased refrigerator-freezers for freezing ice packs, vaccine carriers, cold boxes for transporting vaccines to vaccination sites, and ice packs for maintaining the cold chain.
